# Estimating changes in antibiotic consumption with the introduction of doxycycline post-exposure prophylaxis in the United States

**DOI:** 10.1101/2023.09.20.23295787

**Authors:** Kirstin I. Oliveira Roster, Yonatan H. Grad

**Affiliations:** Department of Immunology and Infectious Diseases, Harvard T.H. Chan School of Public Health, Boston MA

## Abstract

Doxycycline as post-exposure prophylaxis (doxy-PEP) reduces the risk of gonorrhea, chlamydia, and syphilis in studies of men who have sex with men (MSM) and transgender women (TGW) on HIV Pre-exposure Prophylaxis (PrEP) and people living with HIV (PLWH)). Doxy-PEP is an important tool to address the increasing burden of sexually transmitted infections (STIs), but there is concern that increased consumption of doxycycline may drive antimicrobial resistance. We estimated the expected increase in antibiotic use in the US under several doxy-PEP prescribing scenarios. We accounted for doses of antibiotics that may be averted due to the prevention of chlamydia, gonorrhea, and syphilis infections by doxy-PEP. Under a scenario of 75% adoption among the eligible population, with rates of consumption similar to the DoxyPEP trial population, monthly antibiotic consumption would increase by around 2.52 million doses, driven by doxy-PEP consumption of 2.58 million doses and less 62.1 thousand antibiotic doses that would otherwise have been used for chlamydia, gonorrhea, and syphilis treatment.

---

Doxycycline as post-exposure prophylaxis (doxy-PEP), a 200mg dose of doxycycline taken within 72 hours of condomless sex, reduces the risk of gonorrhea, chlamydia, and syphilis in studies of men who have sex with men (MSM) and transgender women (TGW) on HIV Pre-exposure Prophylaxis (PrEP) and people living with HIV (PLWH)) (1,2). Local and state departments of public health have published clinical guidelines for prescribing doxy-PEP (e.g., (3,4)) and national guidelines are expected soon.

Doxy-PEP is an important tool to address the increasing burden of sexually transmitted infections (STIs), but there is concern that increased consumption of doxycycline may drive antimicrobial resistance, including doxycycline resistant *N. gonorrhoeae, Staphylococcus aureus*, and *Streptococcus pneumoniae* (5–8).

How may antibiotic consumption change with the adoption of doxy-PEP? Addressing this question may inform considerations of the risks of antimicrobial resistance and the benefits of STI prevention. Here, we estimated the expected increase in antibiotic consumption in the US under several doxy-PEP prescribing scenarios. We accounted for defined daily doses (DDDs), hereafter simply referred to as *doses*, of antibiotics that may be averted due to the prevention of chlamydia (doxycycline), gonorrhea (ceftriaxone), and syphilis (penicillin) infections by doxy-PEP, with relative risk estimates as reported in the US trial (1) (**Supplemental Table S1**).

We estimated that 0.86 million MSM may be eligible for a doxy-PEP prescription under the enrollment criteria of the DoxyPEP trial (1) (0.53 million PLWH and 0.33 million HIV PrEP users; **Supplemental Table S1**). Under a scenario of 75% adoption among this population (**Supplement**), with rates of consumption similar to the DoxyPEP trial population (4 doses per person-month)(1), monthly antibiotic consumption would increase by around 2.52 million doses, driven by doxy-PEP consumption of 2.58 million doses and less 62.1 thousand antibiotic doses that would otherwise have been used for chlamydia, gonorrhea, and syphilis treatment (**Supplemental Table S2**). Under a scenario of widespread prescribing of doxy-PEP to the entire eligible population (100% adoption), monthly antibiotic consumption would be expected to increase by 3.36 million doses (**Supplemental Table S4**).

A study of ten prescribing strategies based on a patient’s PrEP use, HIV status, and bacterial STI history projected substantial variation across the strategies in the number of infections averted per person taking doxy-PEP (9). The prescribing criterion with the lowest number needed to treat to prevent a chlamydia infection was a diagnosis of two bacterial STIs within six months. Adoption of this strategy with 75% uptake among MSM on HIV PrEP and PLWH would lead to an increase in monthly antibiotic consumption of 0.28 million doses, while widespread (100%) adoption would lead to an increase of 0.37 million doses (**Supplemental Table S5**). Among bacterial STI history-based prescribing strategies, the increase in antibiotic consumption was directly related to the size of the treated population (9); prescribing doxy-PEP to individuals with a diagnosis of at least two concurrent bacterial STIs would be expected to result in the smallest population treated and also the smallest increase in antibiotic consumption (0.18 and 0.24 million doses per month with 75% and 100% adoption, respectively; **Supplement, Equations 2.1-2.3, Table S5**).

A net zero change in doxycycline consumption, where doxy-PEP consumption is balanced with the number of averted doxycycline doses for chlamydia treatment, would require restricting prescriptions to a group with incidence of 7.8 infections per person-year, while maintaining similar levels of monthly doxy-PEP consumption and reductions in chlamydia infection risk reported for HIV PrEP users (**Supplement, Equations 3.1-3.3)**.

Our estimates were based on first-order, short-term use of doxy-PEP. We did not consider the indirect protective effects of doxy-PEP among non-users, which at least initially are expected to reduce bacterial STI incidence (10). We relied on the average estimates of doxy-PEP consumption and relative risk of infection reported in the DoxyPEP trial (1) for all strategies, though these measures likely vary depending on the population treated. Our estimates of antibiotic doses avoided by doxy-PEP considered treatment for uncomplicated chlamydia and gonorrhea and primary syphilis and not for disseminated gonorrhea or for secondary or tertiary disease. Future research is needed to understand consumption and risk among more granular groups and to identify strategies that minimize the number of antibiotic doses consumed (not only the number needed to treat) while preventing the greatest number of STIs.

These estimates suggest that doxycycline consumption is expected to increase with the introduction of doxy-PEP, even when accounting for the reduction in antibiotics used to treat chlamydia, gonorrhea, and syphilis, though the extent of the increase will depend on the size of the population that uses doxy-PEP. Monitoring the changes in the extent of antibiotic consumption, along with changes in disease incidence and the burden of resistance, will be important to understand doxy-PEP’s impact.

## Data Availability

All data are provided in the supplementary materials.

## Funding Statement

This work was supported by National Institute of Allergy and Infectious Diseases (grant numbers R01 AI132606 and R01 AI153521) and CDC contract number 200-2016-91779 to Y. H. G. The findings, conclusions, and views expressed are those of the authors and do not necessarily represent the official position of the Centers for Disease Control and Prevention (CDC).

## Notes

### Competing Interest Statement

The authors have declared no competing interest.

