## Supplement for "Estimating changes in antibiotic consumption with the introduction of doxycycline post-exposure prophylaxis in the United States"

**Supplemental Materials and Methods**

We estimated the net change in monthly antibiotic use, $D_{net}$, as the difference between the number of monthly doses of doxy-PEP consumed by the eligible population, $D_{doxyPEP}$, and the defined daily doses (DDDs) averted, $D_{averted}$, due to reduced chlamydia, gonorrhea, and syphilis infections, ${A= A}_{CT}+ A_{GC}+ A_{S}$, as follows:

$D_{net}=D_{doxyPEP}-A$	(1.1)

$D_{doxyPEP}= \sum_{g} {N_{g} d}_{doxyPEP,g}$	(1.2)

(1.3)

$$D_{averted}=d_{CT}A_{CT}+d_{GC}A_{GC}+d_{S}A_{S}$$

${A= A}_{CT} + A_{GC}+A_{S}= \sum_{c\in\{CT, GC, S\}} \sum_{g} i_{c,g}*\left( 1-RR_{c,g} \right)*N_{g}$	(1.4)

Where:

$D_{doxyPEP}$ *: population-wide doses of doxycycline consumed per month,*

$d_{doxyPEP, g}$*: doses of doxy-PEP consumed per person of group g per month,*

$A$*: doses of antibiotics averted,*

$A_{CT}, A_{GC}, A_{S}$ *: Number of averted cases of chlamydia, gonorrhea, and syphilis, respectively,*

$D_{averted}$ *: Doses of antibiotics averted (in units of defined daily doses, DDD),*

$i_{CT,g}, i_{GC,g}, I_{S,g}$ *: average number of monthly chlamydia, gonorrhea, and syphilis infections per capita in group g,*

$RR_{CT,g}, RR_{GC,g}, RR_{S,g}$ *: relative risk of chlamydia, gonorrhea, and syphilis infection upon adoption of doxy-PEP in group g,*

$d_{CT}, d_{GC}, d_{S}$ *: doses of antibiotics consumed to treat an infection of chlamydia, gonorrhea, and syphilis, respectively,*

$N_{g}$*: population size of group g,*

$g\in\{PrEP, PLWH\}$ *: group, eligible HIV PrEP users or persons living with HIV.*

**Prescribing Strategies**

We considered a range of prescribing scenarios with varying ways of defining the population size of doxy-PEP users. First, we considered different levels of doxy-PEP uptake among the eligible population. Surveys estimated interest in doxy-PEP among MSM with HIV and HIV PrEP users to be 73.5%, and 81.4%, respectively (1). Estimates for likely uptake among the wider population of MSM range from 60% (2) to 84% (3). Based on these figures, we set our uptake parameter to 75% and considered scenarios with 60% and 100% uptake scenarios in sensitivity analyses. Results for these strategies are presented in Tables S2-4.

Secondly, we considered prescribing strategies based on bacterial STI diagnoses, as introduced by Traeger *et al.* (4), ordered by decreasing size of the treated population:

- Strategy #4: Any person with a bacterial STI diagnosis.
- Strategy #5: Any person with a rectal bacterial STI diagnosis.
- Strategy #6: Any person with a gonorrhea diagnosis.
- Strategy #7: Any person with a bacterial STI diagnosis and history of bacterial STI diagnosis within the past 12 months.
- Strategy #8: Any person with a bacterial STI diagnosis and history of bacterial STI diagnosis within the past six months.
- Strategy #9: Any person with a syphilis diagnosis.
- Strategy #10: Any person with diagnosis of at least two concurrent bacterial STIs.

Our calculations are based on the estimates by Traeger *et al.* (4) of the number needed to treat to avert a given infection with chlamydia, gonorrhea, or syphilis. We therefore assume that the magnitude of the benefits of a year-long doxy-PEP prescription scale to the wider population of PLWH and HIV PrEP users, thus also assuming similar levels of incidence among the two groups. We also assumed that the proportion of all MSM PLWH and HIV PrEP users who are treated under each strategy is equal to the proportions given by the Boston community health center cohort in Traeger *et al.* (4). The net increase in consumption of antibiotics under each strategy is presented in Table S5 and was calculated as follows:

$D_{net}=D_{doxyPEP}-A$	(2.1)

$D_{doxyPEP}= \sum_{g} p_{treated,g}*N_{g}* d_{doxyPEP,g}$	(2.2)

${A= A}_{CT} + A_{GC}+A_{S}= \sum_{c\in\{CT, GC, S\}} \sum_{g} p_{treated,g}N_{g}\frac{d_{c}}{12*NNT_{c}}$	(2.3)

Where $p_{treated,g}$ is the proportion of the population of group $g$ treated with doxy-PEP under a given strategy and $NNT_{c}$ is the number needed to treat to avert an infection with condition $c$.

**Identifying a prescribing strategy with a net zero change in antibiotic consumption**

We estimated the chlamydia incidence required among the treated population, such that the net change in doxycycline consumption is zero, meaning the doses of doxy-PEP consumed are balanced by the doxycycline doses averted due to the reduction in chlamydia infections. We used relative risk parameters from the PrEP cohort. We accounted only for chlamydia infections and did not include averted antibiotics from gonorrhea and syphilis cases. Critically, in this estimate, we varied only chlamydia incidence, while keeping doxy-PEP uptake and relative risk estimates fixed. Prescribing doxy-PEP in high-incidence groups would likely lead to either higher doxy-PEP consumption or lower risk reduction, or both, thus biasing our results.

$D_{doxyPEP}=A$	(3.1)

$\sum_{g} d_{doxyPEP,g}N_{g}= \sum_{c\in CT, GC, S\}} d_{c}\sum_{g} i_{c,g}\left( 1-RR_{c,g} \right)N_{g}$	(3.2)

Considering only chlamydia cases averted among a cohort of MSM on HIV PrEP:

$i_{CT,PrEP}=\frac{d_{doxyPEP,PrEP}}{d_{CT}\left( 1-RR_{CT,PrEP} \right)}$	(3.3)

**Notes/Limitations**

We used the same estimates for average monthly use and risk reduction for all strategies, including those based on bacterial STI history. It is very likely that the number of doses consumed to achieve the reductions in infection risk reported in the DoxyPEP trial (5) would differ between the populations treated by those strategies. Antibiotic use may be higher or risk reduction lower among individuals with concurrent or multiple STI diagnoses in the past six or 12 months. More granular estimates of risk reduction and consumption among subsets of MSM on PrEP and PLWH are needed to better anticipate the increase in antibiotic use upon rollout of doxy-PEP and to identify prescribing strategies that most efficiently minimize both the number of doses consumed (not only the number of people needed to treat) together with the number of infections. Understanding how much doxycycline is consumed for each prevented infection will help place the expected increase in selection for resistant strains in context with the reduction in cost and improvement in quality of life expected from reduced bacterial STI transmission.

The above estimates assumed that doxy-PEP consumption is approximately normally distributed and the median and mean monthly doses are approximately equal, meaning that the estimated median consumption in the DoxyPEP trial (5) can be treated as the average consumption per person in our estimates. If consumption follows a long-tailed distribution, with some individuals consuming much more than four doses per month, the mean would be larger than the median, thus raising the estimated monthly consumption of doxy-PEP.

Risk reduction estimates in the DoxyPEP trial were based on quarterly screening visits, which we assumed would be required with doxy-PEP prescriptions during scale-up.

One of the inclusion criteria in the DoxyPEP trial was a history of STI diagnosis in the past 12 months. Since HIV PrEP eligibility has the same requirement, we assumed that all MSM on PrEP would be potentially eligible for doxy-PEP. We further assumed that all MSM with HIV would be eligible. Strict requirement of STI diagnosis in the year prior to doxy-PEP prescription would lower the size of the eligible population of PLWH. Such a reduction in the population size may be treated as equivalent to a scenario with lower uptake, as explored above.

**Table S1. Parameters values**

| **Parameter** | **Description** | **Value** | **Source** |
| --- | --- | --- | --- |
| $d_{g}$ | Median number of doses of doxy-PEP consumed per capita | 4 (IQR 1-10) | (5) |
| $N_{PLWH}$ | US population size of MSM living with HIV | 0.53 million | (6) |
| $N_{PrEP}$ | US population size of people on HIV PrEP | 0.33 million | (7)  Table 9a |
| $i_{CT, PLWH}$ | Chlamydia infection rate among PLWH, mean (95% confidence interval) | 6.0 (5.0 – 7.5) per 100 person-years | (8) |
| $i_{CT,PrEP}$ | Chlamydia infection rate among HIV PrEP users, mean (95% confidence interval) | 38 (20.3 – 55.7) per 100 person-years | (9) |
| $RR_{GC, PLWH}$ | Relative risk of chlamydia infection among PLWH using doxy-PEP (relative to non-users), mean (95% confidence interval) | 0.26 (0.12 - 0.57) | (5) |
| $RR_{GC, PrEP}$ | Relative risk of chlamydia infection among MSM on HIV PrEP using doxy-PEP (relative to non-users), mean (95% confidence interval) | 0.12 (0.05 - 0.25) | (5) |
| $d_{CT}$ | Number of defined daily doses (DDD) of doxycycline averted for each averted chlamydia infection | 7 | (10) |
| $i_{GC, PLWH}$ | Gonorrhea infection rate among PLWH, mean (95% confidence interval) | 6.75 (5.25-8.1)  per 100 person-years | (8) |
| $i_{GC, PrEP}$ | Gonorrhea infection rate among HIV PrEP users, mean (95% confidence interval) | 37.5 (24.3-50.7) per 100 person-years | (9) |
| $RR_{GC, PLWH}$ | Relative risk of gonorrhea infection among PLWH on doxy-PEP (relative to non-users), mean (95% confidence interval) | 0.43 (0.26-0.71) | (5) |
| $RR_{GC, PrEP}$ | Relative risk of gonorrhea infection among MSM on HIV PrEP on doxy-PEP (relative to non-users), mean (95% confidence interval) | 0.45 (0.32-0.65) | (5) |
| $d_{GC}$ | Number of defined daily doses (DDD) of antibiotics averted for each averted gonorrhea infection | 0.25 DDD of ceftriaxone (500mg) | (11) |
| $i_{S, PLWH}$ | Syphilis infection rate among PLWH, mean (95% confidence interval) | 2.5 (2.3 - 2.7) per 100 person-years | (12) |
| $i_{S, PrEP}$ | Syphilis infection rate among HIV PrEP users, mean (95% confidence interval) | 14.5 (3.8-25.2) per 100 person-years | (9) |
| $RR_{S, PLWH}$ | Relative risk of early syphilis infection among PLWH on doxy-PEP (relative to non-users), mean (95% confidence interval) | 0.23 (0.04-1.29) | (5) |
| $RR_{S, PrEP}$ | Relative risk of early syphilis infection among MSM on HIV PrEP on doxy-PEP (relative to non-users), mean (95% confidence interval) | 0.13 (0.03-0.59) | (5) |
| $d_{S}$ | Number of defined daily doses (DDD) of antibiotics averted for each averted early syphilis infection | 2/3 benzathine penicillin G (2.4 million units) | (13) |

**Table S2. Expected increase in monthly defined daily doses (DDDs) of antibiotics consumed under a 75% adoption scenario for different values of doxy-PEP consumption and relative risk of infection**

Estimates are based on 75% uptake among the eligible population; the estimate and upper and lower bounds are defined by the estimates and IQR of monthly doxy-PEP doses consumed as well as the 95% confidence interval in relative risk of chlamydia, gonorrhea, and syphilis infection among doxy-PEP users.

|  |  | Doxy-PEP consumption | | |
| --- | --- | --- | --- | --- |
|  |  | Lower bound  $0.65 milion$ | Estimate  $2.58 million$ | Upper bound  $6.45 million$ |
| Antibiotics averted | Lower bound:  $27.6 thousand$ | $0.62*{10}^{6}$ | $2.55 *{10}^{6}$ | $6.42*{10}^{6}$ |
|  | Estimate: $62.1 thousand$ | $0.58*{10}^{6}$ | $2.52*{10}^{6}$ | $6.39*{10}^{6}$ |
|  | Upper bound: $97.9 thousand$ | $0.55*{10}^{6}$ | $2.48*{10}^{6}$ | $6.35*{10}^{6}$ |

**Table S3. Expected increase in monthly defined daily doses (DDDs) of antibiotics consumed under a 60% adoption scenario for different values of doxy-PEP consumption and relative risk of infection**

Estimates are based on 60% uptake among the eligible population; the estimate and upper and lower bounds are defined by the estimates and IQR of monthly doxy-PEP doses consumed as well as the 95% confidence interval in relative risk of chlamydia, gonorrhea, and syphilis infection among doxy-PEP users.

|  |  | Doxy-PEP consumption | | |
| --- | --- | --- | --- | --- |
|  |  | Lower bound  $0.52 milion$ | Estimate  $2.06 million$ | Upper bound  $5.16 million$ |
| Antibiotics averted | Lower bound:  $22.1 thousand$ | $0.49*{10}^{6}$ | $2.04 *{10}^{6}$ | $5.14*{10}^{6}$ |
|  | Estimate: $49.7 thousand$ | $0.47*{10}^{6}$ | $2.01*{10}^{6}$ | $5.11*{10}^{6}$ |
|  | Upper bound: $78.3 thousand$ | $0.44*{10}^{6}$ | $1.99*{10}^{6}$ | $5.08*{10}^{6}$ |

**Table S4. Expected increase in monthly defined daily doses (DDDs) of antibiotics consumed under a 100% adoption scenario for different values of doxy-PEP consumption and relative risk of infection**

Estimates are based on 100% uptake among the eligible population; the estimate and upper and lower bounds are defined by the IQR of monthly doxy-PEP doses consumed as well as the 95% confidence interval in relative risk of chlamydia, gonorrhea, and syphilis infection among doxy-PEP users.

|  |  | Doxy-PEP consumption | | |
| --- | --- | --- | --- | --- |
|  |  | Lower bound  $0.86*{10}^{6}$ | Estimate  $3.44*{10}^{6}$ | Upper bound  $8.60*{10}^{6}$ |
| Antibiotic doses averted | Lower bound:  $36.8$ *thousand* | $0.82*{10}^{6}$ | $3.40 *{10}^{6}$ | $8.56*{10}^{6}$ |
|  | Estimate:  $82.8 thousand$ | $0.78*{10}^{6}$ | $3.36*{10}^{6}$ | $8.52*{10}^{6}$ |
|  | Upper bound:  $130.5 thousand$ | $0.73*{10}^{6}$ | $3.31*{10}^{6}$ | $8.47*{10}^{6}$ |

**Table S5. Parameters and estimates of doxy-PEP consumption, antibiotics averted, and net change in consumption for different STI history-based prescribing strategies among MSM on HIV PrEP and PLWH**

Estimates are based on average monthly doxy-PEP consumption of 4 doses per person and a population size of N=0.86 million.

| **Strategy** | **Proportion of population treated** | **NNT CT** | **NNT GC** | **NNT syphilis** | **Net increase in consumption (in millions of DDDs per month)** | | |
| --- | --- | --- | --- | --- | --- | --- | --- |
|  |  |  |  |  | **60% uptake** | **75% uptake** | **100% uptake** |
| #4: Any STI | 37.7% | 3.4 | 6.7 | 15.9 | **0.74** | **0.93** | **1.24** |
| #5: rectal STI | 25.4% | 2.9 | 5.9 | 12.5 | **0.50** | **0.62** | **0.83** |
| #6: gonorrhea | 23.2% | 3.0 | 5.9 | 11.6 | **0.45** | **0.57** | **0.76** |
| #7: two STIs in 12 months | 17.1% | 2.5 | 4.7 | 11.0 | **0.33** | **0.41** | **0.55** |
| #8: two STIs in six months | 11.5% | 2.2 | 4.0 | 9.9 | **0.22** | **0.28** | **0.37** |
| #9: syphilis | 8.5% | 3.4 | 5.4 | 10.4 | **0.17** | **0.21** | **0.28** |
| #10: concurrent STIs | 7.6% | 2.2 | 3.7 | 8.6 | **0.15** | **0.18** | **0.24** |
